# Association between peripartum cardiomyopathy and mood disorders in a large, national U.S. cohort

**DOI:** 10.1101/2025.01.02.25319920

**Authors:** Blake Neuburg, Melissa Harris, Anna Palatnik, Rachel Harrison

**Author notes:** **Corresponding Author:** Rachel Harrison, MD Advocate Aurora Health, Downers Grove, IL. Presented at the AHA Hypertension 2023 Scientific Sessions, September 2023, Boston, US. Funding: Dr. Palatnik’s effort is funded by the National Heart Lung and Blood Institute, R34HL165013 and by Eunice Kennedy Shriver National Institute of Child Health and Human Development R01HD112930.

## Abstract

**Objective:** To examine the association between mood disorders in pregnancy and postpartum and peripartum cardiomyopathy (PPCM).

**Methods:** Retrospective cohort study utilizing the National Inpatient Sample from the Healthcare Cost and Utilization Project, Agency for Healthcare Research and Quality of pregnant and postpartum patients from 2017-2019. Patients were separated into two groups based on ICD-10 coding for presence or absence of mood disorder (depression, bipolar depression, anxiety, or other mood diagnosis). The primary outcome was diagnosis of PPCM. Secondary outcomes included a composite of adverse cardiac events and maternal death. Groups were compared via t-tests, chi-squared analysis, and logistic regression that included all variables that differed between groups with p<0.05.

**Results:** Over 2.2 million subjects were analyzed and approximately 168,000 (7.4%) had an ICD-10 diagnosis of mood disorder. Those with mood disorders were more likely to be non-Hispanic white, obese, tobacco users, publicly insured, have comorbidities, and deliver at a large, private, non-profit hospital (all p<0.05). They were delivered at an earlier gestational age and were also more likely to undergo cesarean (37.0±4.6 vs 37.8±3.7 weeks and 35.8% vs 31.8%, respectively)., p<0.001). The primary outcome of PPCM was identified more than twice as often in those with mood disorder (0.12% vs 0.05%, p<0.001). Composite cardiac events, consisting of incidence of acute myocardial infarction, cardiac arrest, cardioversion, cardiac failure, and pulmonary edema were more frequent among patients with mood disorders (0.36% vs 0.14%, p<0.001). After controlling for confounders, PPCM remained independently associated with diagnosis of mood disorders (aOR 1.36, 95%CI 1.03-1.80) as did the composite of adverse cardiac events (aOR 1.57, 95% CI 1.37-1.81).

**Conclusion:** Mood disorders in pregnancy and postpartum are associated with increased rates of PPCM and other cardiac events.

## INTRODUCTION

Peripartum cardiomyopathy (PPCM), defined as a new onset reduced left ventricular ejection fraction of < 45% within the last 3 months of gestation or five months postpartum, is a leading cause of maternal death in the U.S. and worldwide. Peripartum cardiomyopathy attributes to nearly 60% of cardiogenic shock in the peripartum period and often diagnosed in the setting of concomitant pre-eclampsia.^1-4^ The incidence of PPCM diagnosis in the United States is 1 in 1,000 to 4,000 live births and is increasing.^5,6^ Complications of PPCM include severe cardiac morbidity and mortality, which is increasing in incidence from 7.76 to 8.80 per 10,000 over the last 15 years.^7,8^ Various ancestry, biological, and clinical risk factors have been found to be associated with PPCM.^5,6^ There is limited published investigation of the potential impact of mood disorders on the development of PPCM.^3^Further evaluation of those with mood disorders is warranted, not only because mood disorders are associated with hypertensive disorders of pregnancy (HDP) and worse outcomes following cardiovascular events, but also because of the high prevalence of mood disorders.^9,10^

Mental health disorders are common, affecting 1 in 8 to 10 women, and significantly contribute to maternal mortality.^11-13^ Research has shown a strong association between prenatal and postpartum depression and an increased risk of various cardiovascular diseases (CVD), including heart failure, ischemic heart disease, stroke, and chronic hypertension.^9,14-17^ Mood disorders during pregnancy may contribute to this risk through biological mechanisms like HPA axis dysregulation and increased inflammation.^18-20^ Pregnancy acts as a “window period” of vulnerability, potentially heightening women’s susceptibility to CVD and contributing to observed sex disparities in cardiovascular health outcomes.^21-23^ To date, limited research has investigated mood disorders and their association with the development of PPCM. The goal of this study is to test the hypothesis that patients with perinatal mood disorders have higher odds of PPCM and other cardiac events in pregnancy and postpartum.

## METHODS

This was a retrospective cohort study utilizing the Healthcare Cost and Utilization Project National Inpatient Sample (HCUP-NIS), Agency for Healthcare Research and Quality. The HCUP-NIS database is the largest publicly available all-payer inpatient database designed to produce national estimates of inpatient utilization, access, cost, quality and outcomes with a stratified, single cluster sampling design.^31^ Unweighted, it contains approximately 7 million hospital stays annually and weighted designed to approximate a 20% stratified sample representing more than 97% of the United States population per calendar year. We performed analysis on all HCUP-NIS database pregnant and postpartum patients from 2017-2019. Exclusion criteria included male gender, age <11 or >50 years old, and non-pregnant or non-postpartum status. The HCUP-NIS database was queried using International Classification of Diseases-10 (ICD-10) codes to identify a cohort of pregnant or postpartum persons (ICD-10 codes as listed in Supplemental Table 1). Patients with a history of mood disorder (depression, bipolar, anxiety, or other mood diagnosis, ICD-10 codes as listed in Supplemental Table 1) were compared to those without a history of mood disorder. A detailed list of additional ICD-10 codes used is provided in Supplemental Table 1. The primary outcome was diagnosis of PPCM, defined by ICD-10 code. Secondary outcomes included death (as defined by the NIS database) and a composite of acute cardiac events: acute myocardial infarction, cardiac arrest, cardiac conversion, cardiac failure/arrest, and pulmonary edema, defined based on ICD-10 codes (Supplemental Table 1). The following demographic and clinical factors were abstracted from the database and compared between patients with and without mood disorders: maternal age, race and ethnicity, obesity, insurance type, household income quartile, tobacco use, presence of diabetes mellitus, chronic hypertension, asthma, anemia, history of cesarean delivery, limited prenatal care (as ICD coded by provider), multifetal gestation, cesarean delivery, gestational age at delivery, hospital location, type (rural or urban, teaching or non-teaching, private vs. government vs. non-profit), hospital size (based on NIS categorization), and death based on either existing NIS data or ICD-10 codes.

All analyses were performed using Stata (Version 17.0, College Station, TX: StataCorp LLC) Categorical variables were presented as proportions and compared between groups using the Chi-square test. All tests were two sided and p-values < 0.05 were considered statistically significant. The association between mood disorders and PPCM was examined with univariable logistic regression. Adjusted multivariable analysis incorporated the baseline demographic and clinical characteristics that significantly differed between groups with a p<0.05. The secondary outcomes models evaluated maternal death, the cardiac SMM composite outcome and each individual cardiac adverse outcome using univariable and multivariable logistic regressions. Results of multivariable analyses are reported as adjusted odds ratios (aORs) and 95% confidence intervals (CIs). The NIS is supplied as an unidentified, publicly limited data set, as defined by the Health Insurance Portability and Accountability Act, this study was determined to be exempt from review by the Institutional Review Board.

## RESULTS

Over 2.2 million hospitalizations were analyzed and 168,729 (7.4%) included an ICD-10 code diagnosis of mood disorder. Table 1 describes maternal characteristics stratified by presence or absence of a mood disorder. Those with mood disorders were more likely to identify as non-Hispanic white, be obese, use tobacco, have a diagnosis of diabetes mellitus, chronic hypertension, asthma, anemia, be publicly insured, and deliver at a large, private, non-profit hospital (all p<0.001). Patients with mood disorders also were more likely to deliver earlier and undergo cesarean (37.0 ± 4.6 vs 37.8 ± 3.7 weeks and 35.8% vs 31.8%, respectively, p<0.001).

**Table 1.**
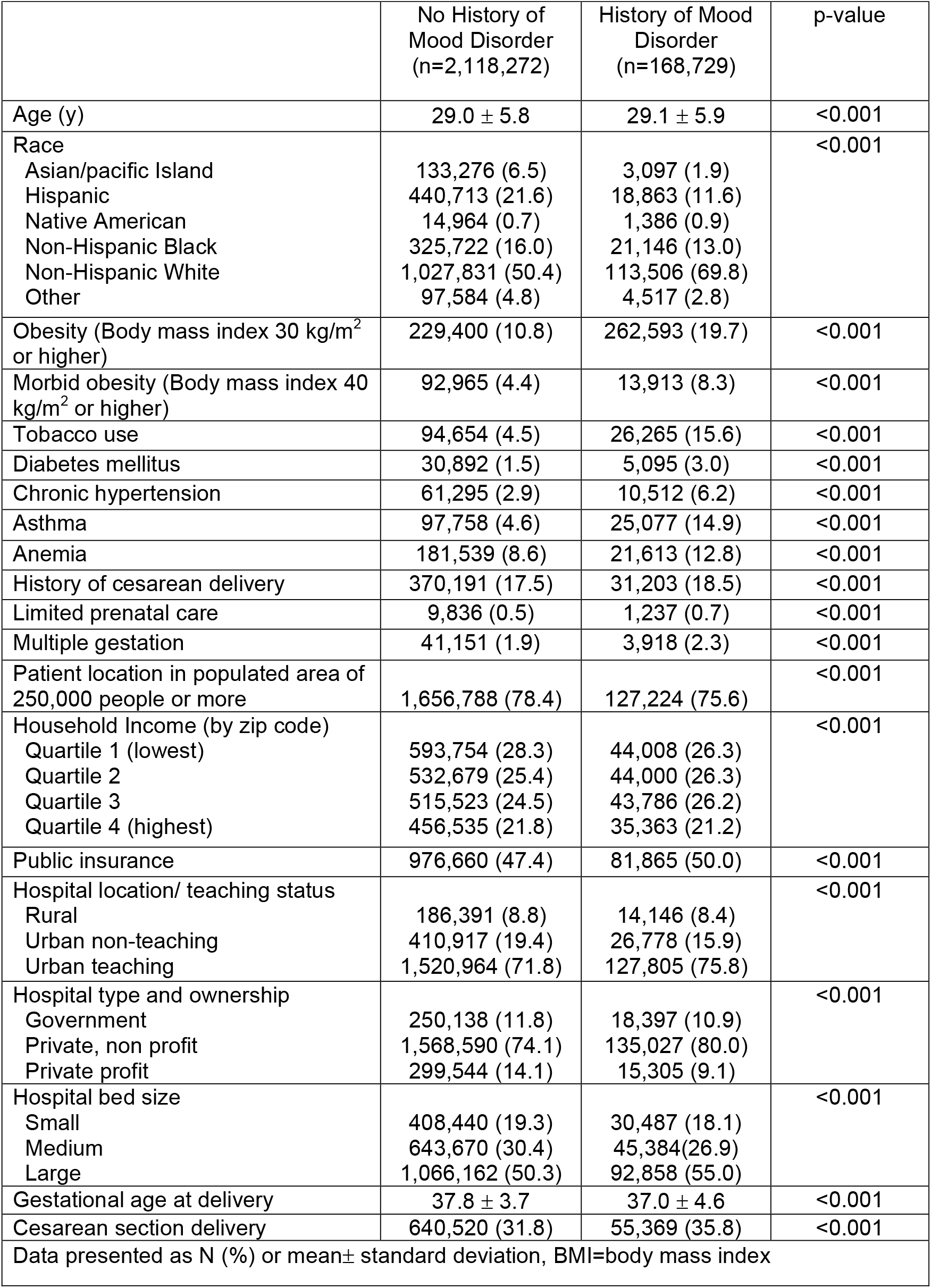
Maternal characteristics stratified by presence or absence of a mood disorder.

Table 2 describes primary and secondary outcomes stratified by presence or absence of a mood disorder. The primary outcome, incidence of PPCM was higher in patients with a mood disorder (0.12% vs 0.05%, p<0.001) as were other cardiac events, including the composite of adverse cardiac events (0.36% vs 0.14%, p<0.001). Each cardiac outcome was statistically significantly associated with mood disorders (p<0.001) except for maternal death (p=0.253) and cardiac failure/arrest as there were too few outcomes to report per NIS reporting guidelines. Table 3 demonstrates the unadjusted and adjusted logistic regressions for the association between PPCM and cardiac morbidity with mood disorders. After adjusting for confounding factors including age, race and ethnicity, obesity, tobacco use, diabetes mellitus, chronic hypertension, asthma, anemia, history of cesarean, limited prenatal care, multiple gestation, location, income, insurance; hospital location, type, ownership, and size PPCM remained independently associated with mood disorders (aOR 1.36, 95% CI 1.03-1.80) as did the composite of adverse cardiac events (aOR 1.57, 95% CI 1.37-1.81) and pulmonary edema (aOR 1.62, 95% CI 1.39-1.89). The other individual components of the composite cardiac morbidity outcome were not associated in the adjusted model.

**Table 2.** Incidence of peripartum cardiomyopathy and cardiac composite outcome stratified by presence or absence of mood disorder.

|  | No History of<br>Mood Disorder<br>(n= 2,118,272) | History of Mood<br>Disorder<br>(n=168,729) | p-value |
| --- | --- | --- | --- |
| Peripartum cardiomyopathy | 1,165 (0.05) | 204 (0.12) | <0.001 |
| Cardiac composite outcome | 2,884 (0.14) | 604 (0.36) | <0.001 |
| Acute myocardial infarction | 153 (0.01) | 40 (0.02) | <0.001 |
| Cardiac arrest | 287 (0.01) | 44 (0.03) | <0.001 |
| Cardiac conversion | 248 (0.01) | 37 (0.02) | <0.001 |
| Cardiac failure/arrest | Too few |  |  |
| Pulmonary edema | 2,453 (0.11) | 519 (0.31) | <0.001 |
| Death | 203 (0.01) | 21 (0.01) | 0.253 |

SMM=severe maternal morbidity,
Cardiac SMM composite includes: acute myocardial infarction, cardiac arrest, cardiac conversion, cardiac failure, pulmonary edema

**Table 3.** Association between peripartum cardiomyopathy and cardiac morbidity by mood disorders.

|  | Unadjusted Odds Ratio | 95% Confidence Interval | Adjusted Odds Ratio* | 95% Confidence Interval |
| --- | --- | --- | --- | --- |
| **Peripartum cardiomyopathy** | **2.20** | **1.90-2.55** | **1.36** | **1.03-1.80** |
| **Cardiac SMM composite outcome** | **2.64** | **2.41-2.88** | **1.57** | **1.37-1.81** |
| Acute MI | 3.28 | 2.32-4.65 | 1.92 | 0.97-3.78 |
| Cardiac arrest | 1.92 | 1.40-2.64 | 1.21 | 0.78-1.88 |
| Cardiac conversion | 1.94 | 1.37-2.74 | 1.29 | 0.82-2.03 |
| Cardiac failure/arrest |  |  |  |  |
| Pulmonary edema | 2.68 | 2.44-2.95 | 1.62 | 1.39-1.89 |
| Death | 1.30 | 0.83-2.04 | 0.64 | 0.32-1.30 |
| SMM=severe maternal morbidity<br>Cardiac SMM composite includes: acute myocardial infarction, cardiac arrest, cardiac conversion, cardiac failure, pulmonary edema<br>*Controlled for: age, race, obesity, tobacco use, diabetes mellitus, chronic hypertension, asthma, anemia, history of cesarean, scant prenatal care, multiple gestation, location, income, insurance; hospital location, type, ownership, and size |  |  |  |  |

## DISCUSSION

In this retrospective cohort study leveraging the NIS database, we identified that perinatal mood disorders were associated with increased rates of PPCM and composite adverse maternal cardiac events. Despite these associations we did not find increased rates of mortality among pregnant individuals with a mood disorder compared to those without.

Previous studies have identified several risk factors for the development of peripartum cardiomyopathy (PPCM), including advanced maternal age, African descent, multifetal pregnancies, and hypertensive disorders of pregnancy.^6,24-26^ The largest of these studies was by Mielniczuk *et al*. examining ∼52 million United States births over a 20-year period from 1990 to 2002.^25^ The authors described increasing incidence of PPCM over the 20-year period from 1 case per 4350 live births in 1990-1993, to 1 case per 2289 live births in 2000-2002 and hypothesized that the primary risk factors driving increasing prevalence were advanced maternal age and increasing rate of multifetal gestations.^25^ Kolte *et al*., published a more recent evaluation of the incidence of PPCM using the NIS database from 2004-2011 and noted continued increasing incidence to ∼1 per 968 live births.^6^ The strongest associations for development of PPCM in that cohort were patients with chronic hypertension, anemia, diabetes, and pre-eclampsia in individuals identifying as African American.^6^ Kolte *et al*. did not assess mood disorders as an independent risk factor specifically but did find higher incidence of depression among women with PPCM during the study period ranging from 5.4 – 7.2%. ^6^ On the other hand, Davis *et al*. published a validated risk predication model for PPCM also found that women with mood disorders were at double the increased risk for PPCM.^3^ Wolfe et al evaluated maternal outcomes among patients with PPCM found women with PPCM had more depressive symptoms and have received medical treatment preconception for mood disorder.^27^ Our study adds to this knowledge by showing that a history of mood disorders is significantly associated with PPCM, with an incidence rate of 1 in 827 among patients with mood disorders. This rate is higher than previously reported, aligns with historical trends, and supports the hypothesis that mood disorders are a significant risk factor for developing PPCM.^3,5,6^

We found an association between mood disorders and other peripartum cardiac events. The predominant diagnosis contributing to cardiac SMM was pulmonary edema. Peripartum and postpartum periods are hallmarked by hemodynamic changes and fluid shifts, which, then coupled with left ventricular diastolic dysfunction of PPCM can often result in pulmonary edema.^28^ Additionally, hypertensive disorders of pregnancy are frequently diagnosed or coincide with PPCM diagnosis and can result in or worsen pulmonary edema.^5,29^

In both obstetric and non-pregnant adult cardiac literature mood disorders are associated with new cardiovascular disease within 24 months postpartum, and a sex specific, independent risk factor for the development of cardiovascular disease.^9,14,30^ Recently a large national cohort study utilizing the Swedish Birth Register examined the association of approximately 878,000 women over a 15 year period, concluding that women who experienced perinatal depression had a 36% higher risk of development of cardiovascular disease.^17^ Among the cohort, those with history of psychiatric disorders had the highest association for the development of cardiovascular disease and the association was significant for all cardiovascular disease subtypes including ischemic heart disease and heart failure. While cardiovascular disease is the leading cause of death in the general population, individuals with significant mental health disorders are disproportionally affected by cardiovascular disease.^31,32^ A large metanalysis examining the incidence of cardiovascular disease and mortality in over 3.2 million patients with significant mental illness compared with 113 million controls found individuals with significant mental illness had an approximately odds ratio of 1.53 increased risk of cardiovascular disease relative to controls across the 11 studies included in the analysis.^33^

Our study findings suggest that further investigation is warranted to further characterize the relationship between mood disorders and peripartum cardiomyopathy.^31^ There are significant ongoing efforts to identify causes and reduce increasing rates of maternal mortality in the United States and peripartum cardiomyopathy is a leading cause of maternal mortality.^1,34^ Current proposed prediction models to identify women at risk for the development of PPCM have identified mood disorders as a risk factor.^3^ Pregnant individuals who are identified to be at risk of development for PPCM who have a mood disorder could be referred for mental health services and optimization while individuals who develop PPCM could be screened for the development of mood disorders. Whether the treatment of mood disorders can affect pregnant patients recovery from PPCM and long-term cardiovascular health requires further study.^7,17^

Our study has several strengths. It included a large sample size that allowed us to study rare outcomes such as PPCM and other cardiac events. Furthermore, the association of PPCM with mood disorders identified a modifiable risk factor and presents opportunities to prevent or improve outcomes for patients with PPCM.^35^ Limitations of our study include the use of ICD-10 codes for research purpose however, prior studies have demonstrated reasonable accuracy with their use.^6^ Additional limitations to any study utilizing the NIS dataset are that it can only assess inpatient hospitalizations, thus, the time course for events makes it difficult to elicit causation versus association. The NIS does not report post discharge data and thus long-term outcome data is absent.

## CONCLUSIONS

In conclusion, we found that perinatal mood disorders were associated with increased rates of PPCM and other cardiac events. These findings have implications for mental health care in pregnancy and postpartum and the prevention of PPCM and adverse cardiac events.

## Data Availability

All data used in study was access as part of the National (Nationwide) Inpatient Sample (NIS) database for for years 2017-2019

https://hcup-us.ahrq.gov/nisoverview.jsp

## Acknowledgements

National Inpatient Sample data partners as listed here: https://hcup-us.ahrq.gov/db/hcupdatapartners.jsp

---

**Supplemental Table 1.** ICD-10 or procedure codes.

|  |  |
| --- | --- |
| Depression | F33.0-F33.9 F32.0-F32.9 |
| Bipolar disorder | F30.0-F30.9 F31.0-F31.9 |
| Postpartum depression | F53.0 |
| Anxiety | F41.0-F41.9 |
| Mood – other | F34.0-F34.9 F39.0-F39.9 |
| Obesity (body mass index 30.0 kg/m<sup>2</sup> or greater) | O99.210-O99.215 E66.0-E66.2 E66.8-E66.9 Z68.3 Z68.4 Z68.30-Z68.99 |
| Tobacco use | F17.200 F17.203-F17.209 F17.210 F17.213-F17.219 F17.220 F17.223-F17.229 F17.290 F17.293-F17.299 O99.330-O99.335 |
| Diabetes mellitus | O24.0-O24.1 E10.00-E10.90 O24.8-O24.93 E11.00-E11.90 |
| Chronic hypertension | O10.0-O10.93 I10.0 I15.0-I15.9 O16.1 |
| Asthma | J45.2-J45.998 O99.5 O99.519 |
| Anemia | O90.81 O99.0-O99.019 D50.9 D64.9-D64.901 |
| History of cesarean | O34.2-O34.22 O66.41 |
| Scant prenatal care | O09.30-O09.33 |
| Multiple gestation | O30.0-O30.93 |
| Cesarean delivery | 10D00Z0-10D00Z2 10A00ZZ 10A03ZZ 10Z04ZZ |
| Peripartum cardiomyopathy | O90.3 |
| Acute myocardial infarction | I21.01-I21.9 I21.A1 I21.A9 I22.0-I22.9 I23.0-I23.8 I97.71 |
| Cardiac arrest | I46.2 I46.8 I46.9 I49.01 I49.02 |
| Cardiac conversion | 5A2204Z 5A12012 |
| Cardiac failure/arrest | I97.120-I97.131 I97.710 I97.711 |
| Pulmonary edema | J81.0 I50.1 I50.20 I50.21 I50.23 I50.30 I50.31 I50.33 I50.40 I50.41 I50.43 I50.9 J68.1 |
| Pregnancy or postpartum | O0.0-O99.99 Z39.0-Z39.2 |

